# “Efficacy and Safety of Fixed Combination of Hydroxychloroquine with Azithromycin Versus Hydroxychloroquine and Placebo in Patients with Mild COVID-19: Randomized, double blind, Placebo controlled trial”

**DOI:** 10.1101/2022.04.06.22273531

**Authors:** Ivonne A Roy-García, Moises Moreno-Noguez, Rodolfo Rivas-Ruiz, Marta Zapata-Tarres, Marcela Perez-Rodriguez, Magaly A Ortiz-Zamora, Lourdes Gabriela Navarro-Susano, Lilia M Guzman-Rivas, Luis Rey Garcia-Cortes, Icela Palma-Lara, Pedro Gutierrez-Catrellón

## Abstract

To determine the efficacy and safety of fixed combination of hydroxychloroquine/azithromycin (HCQ+AZT) compared to hydroxychloroquine (HCQ) alone or placebo in mild COVID-19 outpatients to avoid hospitalization.

**Materials and methods:** This randomized, parallel, double-blind clinical trial included male and female patients aged 18 and 76 years non COVID vaccinated, who were diagnosed with mild COVID-19 infection. All patients underwent liver and kidney profile test, as well as a health questionnaire and clinical revision to document that they did not have uncontrolled comorbidities. They were randomly assigned to one of the three treatment arms: 1) hydroxychloroquine with azithromycin 200 mg/250 mg every 12 hours for five days followed by hydroxychloroquine 200 mg every 12 hours for 5 days; 2) hydroxychloroquine 200 mg every 12 hours for ten days; or 3) placebo every 12 hours for ten days. The primary outcome of the study was hospitalization, while the secondary outcomes were disease progression, pneumonia, use of supplemental oxygen, and adverse events. This study was registered in clinicaltrials.gov with the NCT number of 04964583.

**Results:** A total of 92 participants were randomized. Of whom, 30 received HCQ+AZT, 31 received HCQ, and 31 received placebo. The median age was 37 years, 27.2% of the participants had comorbidities, and the global incidence of hospitalization was 2.2%. The incidence of hospitalization was 6.7% (2/30) in the HCQ+AZT group compared to the HCQ or placebo groups, in which there were no hospitalizations. Progression of disease was higher in the HCQ group [RR=3.25 (95% CI, 1.19-8.87)] compared with placebo group. There was no statistical difference between the HCQ+AZT group and the placebo group in progression of disease. The incidence of pneumonia was 30% in the HCQ+AZT group, 32.2% in the HCQ group, and 9.6% in the placebo group (HCQ + AZT vs Placebo; p=0.06). There was a significant risk of pneumonia versus placebo only in the HCQ group [RR=3.33 (95% CI, 1.01-10.9)]. Supplemental oxygen was required by 20% (6/30) of the patients in the HCQ+AZT group, 6.4 (2/31) of the patients in the HCQ group, and 3.2% (1/31) of the patients in the placebo group,[(HCQ + AZT vs Placebo; p=0.100), (HCQ vs Placebo, p=0.610)]. There was no statistical difference between groups for negative test (PCR) on day 11. The most frequent adverse events were gastrointestinal symptoms. No lengthening of the QT interval was observed in patients receiving HCQ+AZT or HCQ.

**Conclusion:** The use of HCQ+AZT does not decrease the risk of hospitalization in patients with mild COVID-19. The use of HCQ increases the risk of progression and pneumonia.

## Introduction

At the end of December 2019, a new viral infection belonging to the Coronavirus family emerged in China [1], which spread rapidly worldwide [2] and was declared a pandemic by the WHO in March 2020. The COVID-19 disease requires hospitalization in 20% of patients, 33.7%of whom require admission to the intensive care unit, with a mortality rate reaching up to 62.4% [3]. The need to find an urgent treatment to reduce the impact of the pandemic has led to the use of different therapeutic options, among them is the fixed combination of hydroxychloroquine and azithromycin which became a promising option [4,5].

Hydroxychloroquine has antiviral activity, inhibits endosome acidification, interferes with virus fusion to the cell, exhibits non-specific antiviral activity in vitro against a wide range of emerging viruses (HIV, dengue, hepatitis C, SARS, and MERS), and more recently against SARS CoV-2 in addition to its anti-inflammatory activity [6,7]. On the other hand, azithromycin is a macrolide antibiotic indicated for airway infections, which has shown anti-inflammatory and antiviral effects in vitro [8], which is why it was proposed as an accessible and cost-effective option for SARS CoV-2 infection [9].

Even if these drugs have demonstrated efficacy in vitro nowadays, clinical results on the beneficial effect of the fixed combination HCQ+AZT in hospitalized or mechanically ventilated patients are inconsistent [10,11], which is attributed to the delayed initiation of treatment when viral replication and inflammatory response characterized by an increased cytokine storm have already occurred [12]. Therefore, it is hypothesized that early use of these drugs is necessary to achieve benefits to prevent hospitalizations. This study aimed at determining the efficacy and safety of the fixed combination of HCQ+AZT compared to HCQ or placebo in mild COVID-19 outpatients to avoid hospitalizations [13].

## Methods

### Trial design

This study was designed as a multicenter, parallel, double-blind, randomized clinical trial. The study was conducted in two public ambulatory family medicine units, in Mexico City (UMF No. 28) and at the State of México (UMF No. 52) both of the Instituto Mexicano del Seguro Social (IMSS), which is the national social security institute. The trial was designed and conducted by the authors. The protocol was approved by the National Research Committee institution’s review board of the IMSS with the number R-2020-785-138 and by COFEPRIS, the Mexican drug regulatory agency. Written informed consent was obtained from each of the included participants. Ultra laboratories were the sponsor of the study; nonetheless, they were not involved in protocol or the analysis of results or in the preparation of the manuscript.

The trial was registered in clinicatrials.gov (Clinical Trials: NCT04964583). This report follows the CONSORT guidelines.

### Eligible subjects

The study included patients aged 18-76 years who were diagnosed with mild COVID-19 with acute respiratory disease and who met the current operational definition of the Ministry of Health of Mexico, which included the presence of a major symptom such as headache, fever, cough, dyspnea, and any of the following minor symptoms: myalgia, arthralgia, odynophagia, chills, chest pain, rhinorrhea, anosmia, dysgeusia, or conjunctivitis [14]. The diagnosis was confirmed by RT-PCR for SARS-CoV-2, the severity of the disease was evaluated with the NEWS scale, patients with a score ≤4 points were considered to have mild disease. Patients with cardiac disorders with delayed cardiac conduction (QT segment ≥ 450 ms), pregnant or lactating women, patients with hypersensitivity to study drugs, patients with chronic renal failure with (eGFR<40 mL/min), patients with a history of retinopathy or macular degeneration, known Glucose 6 Phosphate Dehydrogenase (G6PD) deficiency, patients with liver disease, cirrhosis or those using the following medications: colchicine, ergotamine, dihydroergotamine, citalopram, hydroxyzine, domperidone, piperazine, antiarrhythmic drugs class IA and III and antidepressant medications were excluded from the study.

### Intervention

Participants were randomly assigned to one of the three treatment arms. The participants in Group 1 (HCQ+AZT) received hydroxychloroquine 200 mg/azithromycin 250 mg orally every 12 hours for 5 days, followed by hydroxychloroquine 200 mg every 12 hours for 5 more days. The participants in Group 2 (HCQ) received only hydroxychloroquine 200 mg orally every 12 hours for 10 days. The participants in Group 3 received an oral placebo every 12 hours for 10 days. All three treatment groups received symptomatic treatment for the management of COVID-19, the current standard was at discretion of the treating physician based on acetaminophen, non-steroidal analgesics, antihistamines and Ivermectin. A computerized random number sequence was generated for assignment to study treatment groups and was stratified according to center. Randomization was carried out by one of the investigators who did not participate in the inclusion of patients or in the delivery of medication. The medical staff was responsible for the recruitment and selection of participants and the evaluation of the correct allocation of treatment according to the randomization. Adherence to treatment was evaluated by counting the tablets during the medical consultation and by the intake recorded by the participants in the digital application for smartphones.

### Follow-up

The follow-up was carried out through a Web App on a smartphone explicitly elaborated for the research project (digital monitoring), for which the participant was previously instructed, recording the medication intake, adverse events, oxygen saturation, and temperature during the ten days of the treatment. A clinical examination was performed at the time of inclusion, on days 6 and 11 after initiating the treatment, in which an evaluation of the severity of the disease was carried out along with an RT-PCR test, electrocardiogram and follow-up laboratory studies, count of tablets to evaluate therapeutic adherence, and radiography taken at the beginning of the study and on day 11. On day 21 of the follow-up, the participants were called by phone to evaluate symptoms and to investigate whether the patient had returned to routine activities, required hospitalization, or had any of the secondary outcomes. All participants who showed a change suggestive of deterioration (oxygen saturation less than 90% in room air, and shortness of breath or pneumonia) during their digital follow-ups were contacted by a specialized physician to confirm the data and establish the behavior to follow.

### Outcomes

The primary outcome of the study was hospitalization during the 21 days of the follow-up. The secondary outcomes included disease progression, which was defined by oxygen saturation less than 90%, dyspnea, or pneumonia [15], use of supplemental oxygen, the presence of adverse events during the 21 days of follow-up, and PCR results on days 6 and 11 of the follow-up.

### Drug Safety

All adverse events reported by the participants during the study period in the electronic patient diary app or during medical consultations were recorded. To assess the safety of the drug, QT intervals were measured by a certified cardiologist using the Bazett formula by performing a 12-lead electrocardiogram at the inclusion visit, on days 6 and 11 of the follow-up.

## Sample Size

The sample size was calculated using a comparative study formula. An expected difference of 40% in the clinical response between HCQ+AZT treatment and placebo was considered, with a margin of superiority of 10%, an α error of 5%, and a statistical power of 80%. The estimated sample size obtained from the calculation was 84 participants, 28 participants per group, to which 20% was added for possible losses, obtaining a sample size of 105 participants. Due to the decrease in the number of COVID-19 patients, only 92 participants were included.

### Statistical Analysis

#### Therapeutic Efficacy Analysis

The therapeutic efficacy was assessed using an intention-to-treat analysis. The variables considered for the determination of efficacy were as follows: hospitalization, disease progression, pneumonia, and use of supplemental oxygen.

#### Clinical Safety Analysis

The safety analysis was carried out considering all the participants regardless of their completion of the study. The frequency and severity of the adverse events presented by the participants during the study were determined, and a comparison was made according to the types of treatment given using the Pearson’s X^2^test or Fischer exact test.

#### Descriptive and Inferential Statistics

A descriptive analysis was carried out to determine the general characteristics of the population [16].

To determine if there were differences in the baseline status of the population by the treatment assignment, the ANOVA test was used for systolic and diastolic pressure, PCR (CT), total leukocytes and lymphocytes, the Kruskal-Wallis test was used for age, BMI, temperature, oxygen saturation, NEWS Score, Glucose, urea, lactic dehydrogenase, GGT, transaminases, serum iron, neutrophils and platelets, and the Pearson’s X^2^ test or Fisher’s exact test was used for sex, comorbidities, smoking and Ivermectin.

The incidence of hospitalization, disease progression, pneumonia, use of supplemental oxygen and negative test (PCR) on day 11 was determined according to the treatment assignment as a measure of association. The relative risk (RR) was calculated, with its 95% confidence interval, and the number needed to treat (NNT) and number needed to harm (NNH) were calculated as a measure of potential impact [17].

Alpha was set at 5%, all data were analyzed by using IBM SPSS v.28 (IBM Corp, NY, USA).

## Results

### Study population

Figure 1 shows the number of participants included in the study and assigned to treatments. A total of 92 participants were randomized between January and June 2021 and included in the intention-to-treat analysis. In the per-protocol analysis, 6 participants were excluded, four patients for withdrawing informed consent in the HCQ+AZT group and one for medication error, and one participant for withdrawing informed consent in the HCQ group. None of the patients was excluded in the placebo group.

**Figure 1.**
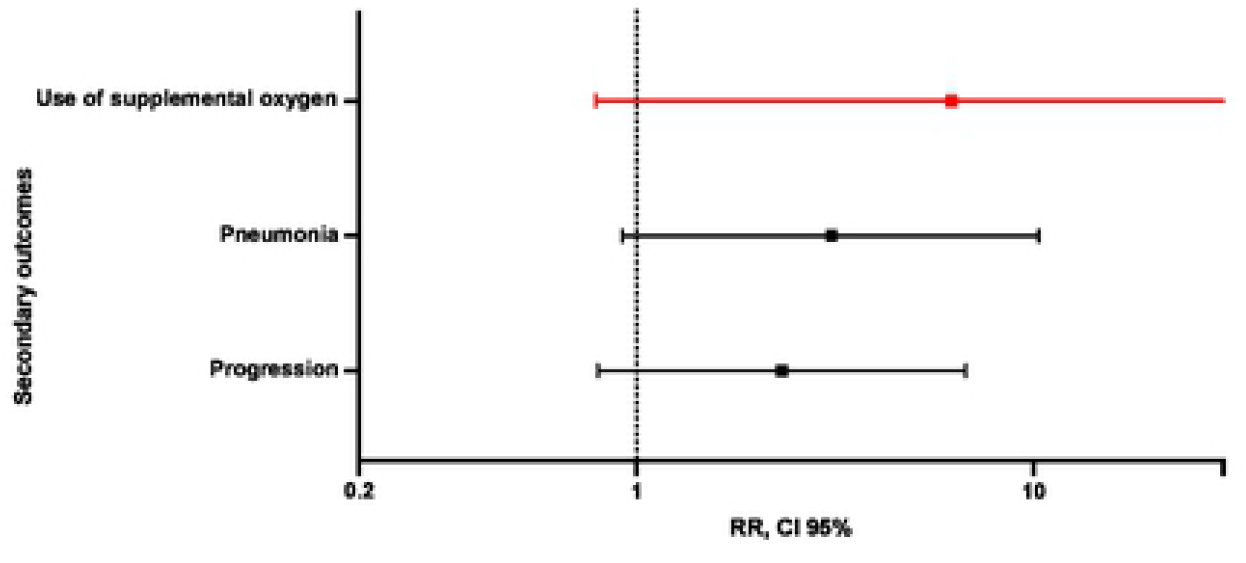
Efficacy of HCQ+AZT vs placebo for secondary outcomes.

**Figure 2.**
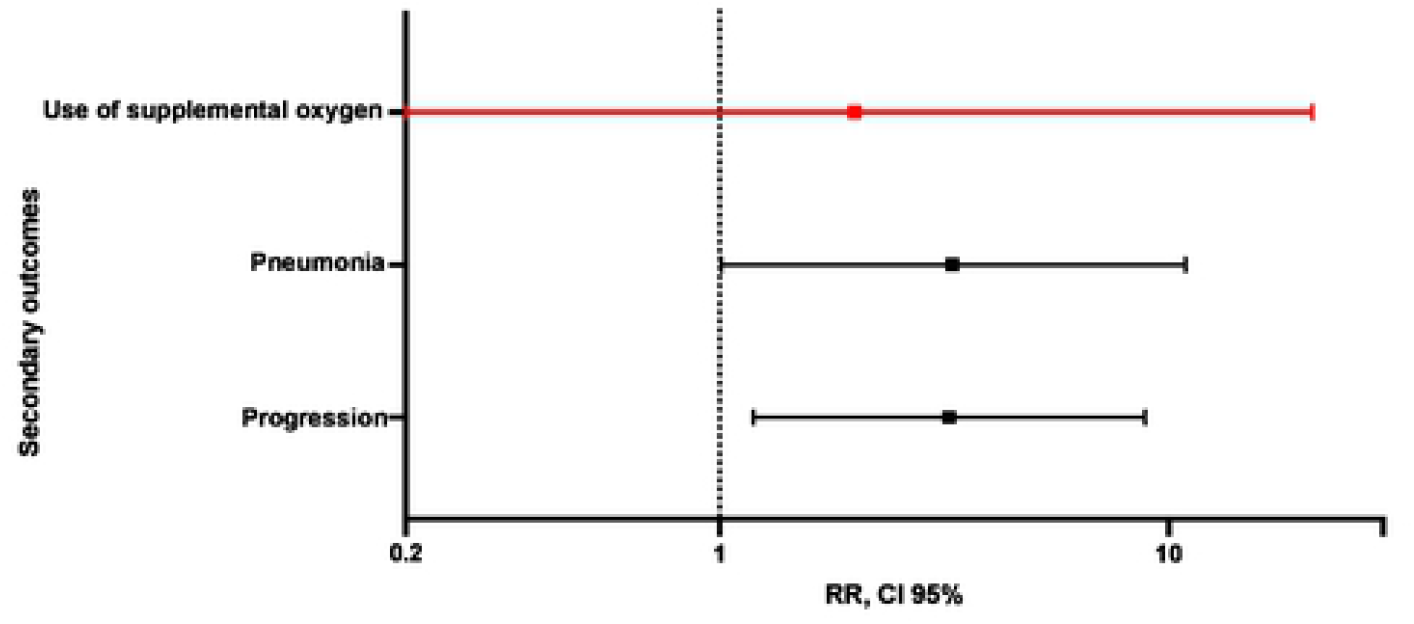
Efficacy of HCQ vs placebo for secondary outcomes.

The general characteristics of the population are shown in Table 1. The median age was 37 years, 27.2% of the patients had comorbidities, the median BMI was 26.9. The analysis of the time of evolution at the initiation of treatment revealed a median of 5 days, with a median saturation of 94%. The incidence of hospitalization in the study population was 2.2%. The analysis of the laboratory studies revealed that 27.2% of the participants had elevated lactic dehydrogenase levels, 45.7% had elevated gamma-glutamyl transferase levels, 48.9% had elevated levels of some type of transaminase, 8.7% had neutropenia, and 21.7% had thrombocytopenia at the beginning of the study.

**Table 1.**
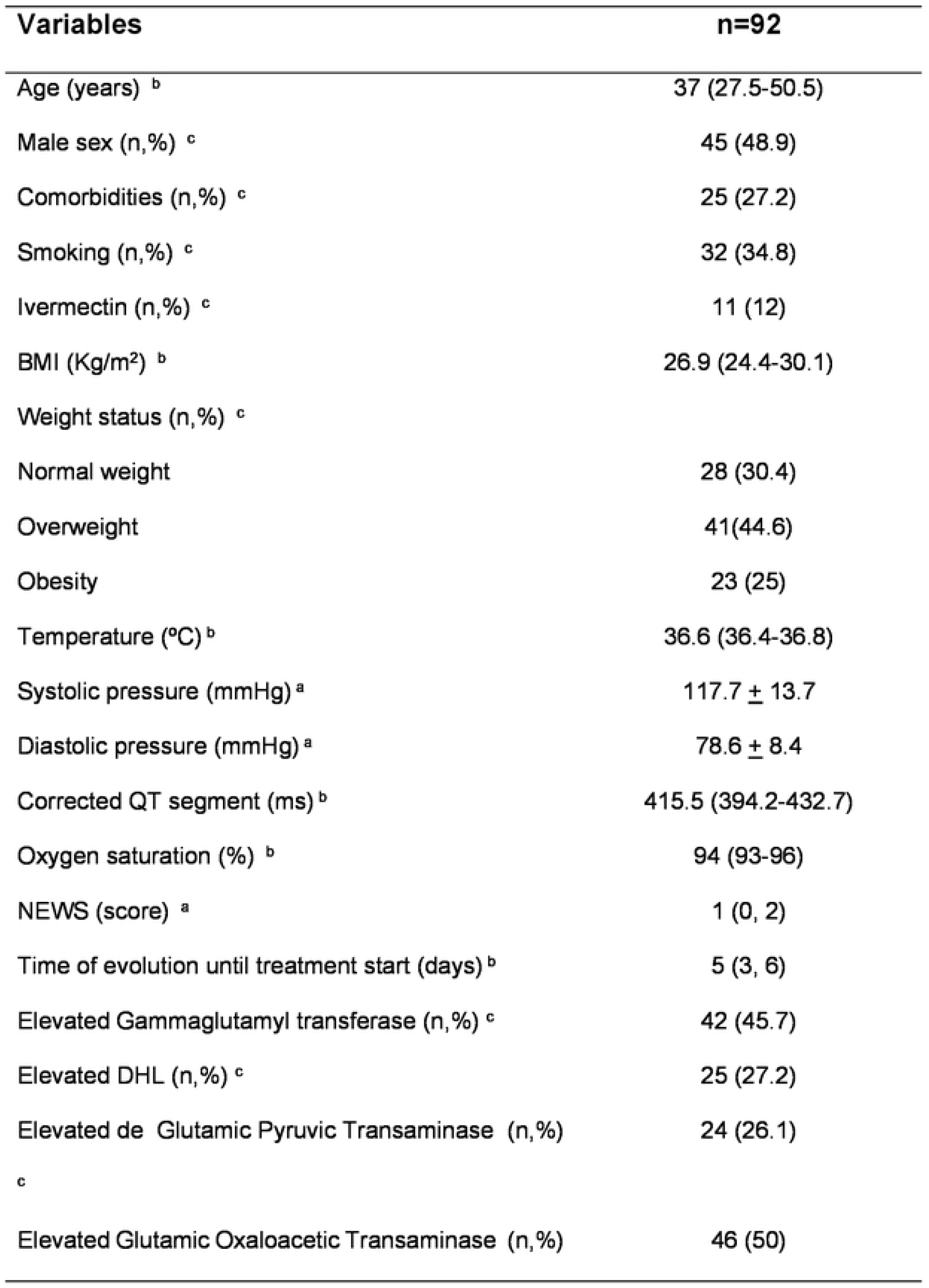
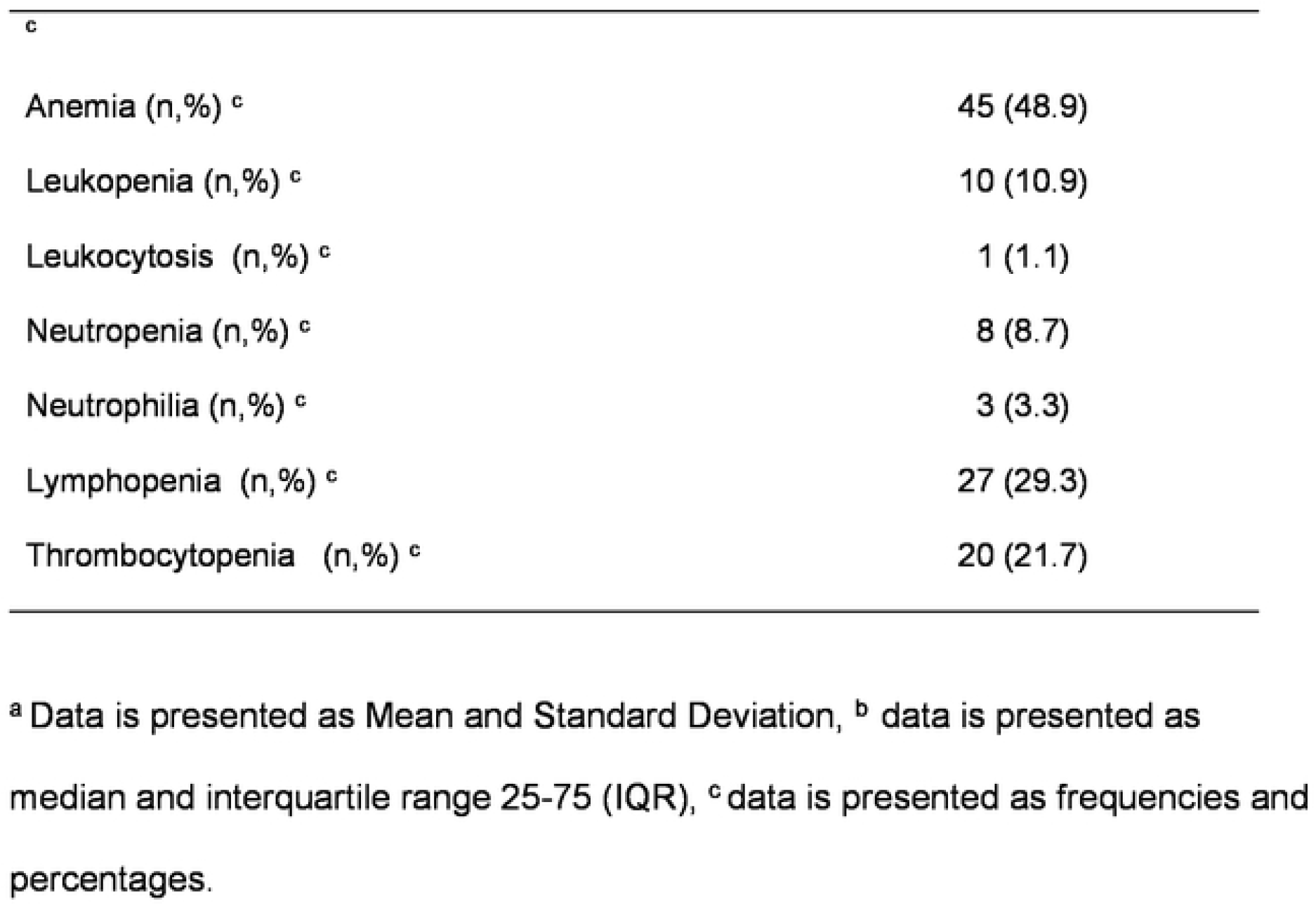
Baseline characteristics of participants

| **Variables** | **n=92** |
| --- | --- |
| Age (years) <sup>b</sup> | 37 (27.5-50.5) |
| Male sex (n,%) <sup>c</sup> | 45 (48.9) |
| Comorbidities (n,%) <sup>c</sup> | 25 (27.2) |
| Smoking (n,%) <sup>c</sup> | 32 (34.8) |
| Ivermectin (n,%) <sup>c</sup> | 11 (12) |
| BMI (Kg/m<sup>2</sup>) <sup>b</sup> | 26.9 (24.4-30.1) |
| Weight status (n,%) <sup>c</sup> |  |
| Normal weight | 28 (30.4) |
| Overweight | 41(44.6) |
| Obesity | 23 (25) |
| Temperature (°C) <sup>b</sup> | 36.6 (36.4-36.8) |
| Systolic pressure (mmHg) <sup>a</sup> | 117.7 ± 13.7 |
| Diastolic pressure (mmHg) <sup>a</sup> | 78.6 ± 8.4 |
| Corrected QT segment (ms) <sup>b</sup> | 415.5 (394.2-432.7) |
| Oxygen saturation (%) <sup>b</sup> | 94 (93-96) |
| NEWS (score) <sup>a</sup> | 1 (0, 2) |
| Time of evolution until treatment start (days) <sup>b</sup> | 5 (3, 6) |
| Elevated Gammaglutamyl transferase (n,%) <sup>c</sup> | 42 (45.7) |
| Elevated DHL (n,%) <sup>c</sup> | 25 (27.2) |
| Elevated de Glutamic Pyruvic Transaminase (n,%) | 24 (26.1) |
| <sup>c</sup> |  |
| Elevated Glutamic Oxaloacetic Transaminase (n,%) | 46 (50) |
| <sup>c</sup> |  |
| Anemia (n,%) <sup>c</sup> | 45 (48.9) |
| Leukopenia (n,%) <sup>c</sup> | 10 (10.9) |
| Leukocytosis (n,%) <sup>c</sup> | 1 (1.1) |
| Neutropenia (n,%) <sup>c</sup> | 8 (8.7) |
| Neutrophilia (n,%) <sup>c</sup> | 3 (3.3) |
| Lymphopenia (n,%) <sup>c</sup> | 27 (29.3) |
| Thrombocytopenia (n,%) <sup>c</sup> | 20 (21.7) |

<sup>a</sup> Data is presented as Mean and Standard Deviation, <sup>b</sup> data is presented as median and interquartile range 25-75 (IQR), <sup>c</sup> data is presented as frequencies and percentages.

The analysis of the symptoms at the beginning of the study showed that 81.5% of the participants had a cough, 77.2% had a headache, 62% had fatigue, 32.6% had a fever, 23.9% had arthralgia, 8.7% had dyspnea, 23.9% had a sore throat, 32.6% had anosmia, and 30.4% had dysgeusia.

Table 2 shows the baseline characteristics of the population according to the allocation to the treatment arm. There were no significant differences for any of the variables analyzed.

**Table 2.** General characteristics according to treatment arm

| Variables | Total | HCQ | HCQ+AZT | Placebo | p |
| --- | --- | --- | --- | --- | --- |
|  | n= 92 | n=31 | n=30 | n=31 |  |
| Age (years) <sup>b</sup> | 37 (27.5-50.5) | 38 (24-49.5) | 41 (33.7-49.5) | 32 (24-51) | 0.34 |
| Male sex (n,%) <sup>c</sup> | 45 (48.9) | 13 (41.9) | 12 (40.0) | 20 (64.5) | 0.07 |
| Comorbidities (n,%) <sup>c</sup> | 25 (27.2) | 9 (29.0) | 7 (23.3) | 9 (29.0) | 1.0 |
| Smoking (n,%) <sup>c</sup> | 32 (34.8) | 10 (32.3) | 9 (30.0) | 13 (41.9) | 0.42 |
| Time of evolution until treatment start<br>(days) <sup>b</sup> | 5 (3, 6) | 5 (4, 6) | 5 (3, 6) | 5 (3, 7) | 0.78 |
| BMI (Kg/m<sup>2</sup>) <sup>b</sup> | 26.9 (24.4- 30.1) | 26.9 (24.6-28.6) | 27.3 (25.2-30.4) | 25.6 (24.1-31.7) | 0.53 |
| Temperature (°C) <sup>b</sup> | 36.6 (36.4-36.8) | 36.7 (36.4-36.8) | 36.6 (36.4-36.8) | 36.6 (36.5- 36.8) | 0.97 |
| Systolic pressure (mmHg) <sup>a</sup> | 117.7 ± 13.7 | 117.2 ± 13.9 | 117.0 ± 12.5 | 118.3 ± 14.7 | 0.93 |
| Diastolic pressure (mmHg) <sup>a</sup> | 78.6 ± 8.4 | 78.3 ± 8.1 | 78.5 ± 8.8 | 78.9 ± 8.8 | 0.97 |
| Oxygen saturation (%) <sup>b</sup> | 94 (93-96) | 95 (93.7-96) | 94 (92-95) | 94 (93-96) | 0.53 |
| NEWS (score) <sup>b</sup> | 1 (0,2) | 0.5 (0, 2) | 1 (0, 2) | 2 (0, 2) | 0.43 |
| PCR (CT) <sup>a</sup> | 26.6 ± 4.7 | 26.2 ± 4.8 | 26.4 ± 4.9 | 27.1 ± 4.5 | 0.72 |
| Glucose (mg/dL) <sup>b</sup> | 88.7 (79.3-96.3) | 92.4 (75.5-101.6) | 88.2 (78.8-99.5) | 84.6 (79.6-92.5) | 0.28 |
| Urea (mg/dL) <sup>a</sup> | 25.5 (21.1-33.1) | 26.8 (21.0-33.6) | 25.1 (19.5-33.3) | 24.9 (22.1-32.8) | 0.85 |
| Lactic dehydrogenase (U/L) <sup>b</sup> | 196.6 (164.6-221.1) | 198.3 (158.4-231.4) | 194.5 (167.7-217.5) | 182.9 (166.5-220.6) | 0.97 |
| Gamma Glutamyl Transferasa (U/L) <sup>b</sup> | 32.9 (21.1-66.1) | 31.8 (21.2-66.9) | 35.8 (19.2- 73.3) | 32.8 (21.9-65.7) | 0.98 |
| Glutamic Pyruvic Transaminase (U/L) <sup>b</sup> | 33.6 (19.9- 55.3) | 36.7 (21.6- 54.1) | 33.4 (23.3- 58.7) | 24.2 (15.4-54.3) | 0.45 |
| Glutamic Oxaloacetic Transaminase (U/L) <sup>b</sup> | 31.5 (25.3- 44.5) | 32.1 (27.7- 46.8) | 32.0 (24.7- 42.1) | 29.6 (23.7- 40.8) | 0.53 |
| Serum iron (ug/dL) <sup>a</sup> | 50.8 (35.4-69.4) | 48.5 (32.7- 69.8) | 55.1 (37.6- 73.2) | 47.2 (35.3-65.3) | 0.56 |
| Total leukocytes (K/μL) <sup>a</sup> | 4.99 ± 1.49 | 5.1 ± 1.5 | 5.1 ± 1.6 | 4.8 ± 1.2 | 0.78 |
| Neutrophils (K/μL) <sup>b</sup> | 2.7 (2.3-3.8) | 2.7 (2.3-4.1) | 2.8 (2.1- 3.9) | 2.7 (1.9-3.7) | 0.71 |
| Lymphocytes (K/μL) <sup>a</sup> | 1.2 ± 0.4 | 1.2 ± 0.4 | 1.2 ± 0.4 | 1.2 ± 0.4 | 0.99 |
| Platelets (K/μL) <sup>b</sup> | 212 (172-256.5) | 212 (175-257) | 202 (157.7- 266) | 218.5 (181-257.5) | 0.76 |

<sup>a</sup> Data is presented as Mean, Standard Deviation and statistical was done was ANOVA, <sup>b</sup> data is presented as median and interquartile range 25-75 (IQR), statistical test was Kruskal Wallis, <sup>c</sup> data is presented as frequencies and percentages, statistical test was X<sup>2</sup>

### Primary Outcome

The analysis of the therapeutic efficacy to reduce the risk of hospitalization revealed that the incidence of the outcome in the HCQ+AZT group was 6.7% (2/30) compared to the HCQ group and the placebo group, in which there were no hospitalizations (Table 3).

**Table 3.**
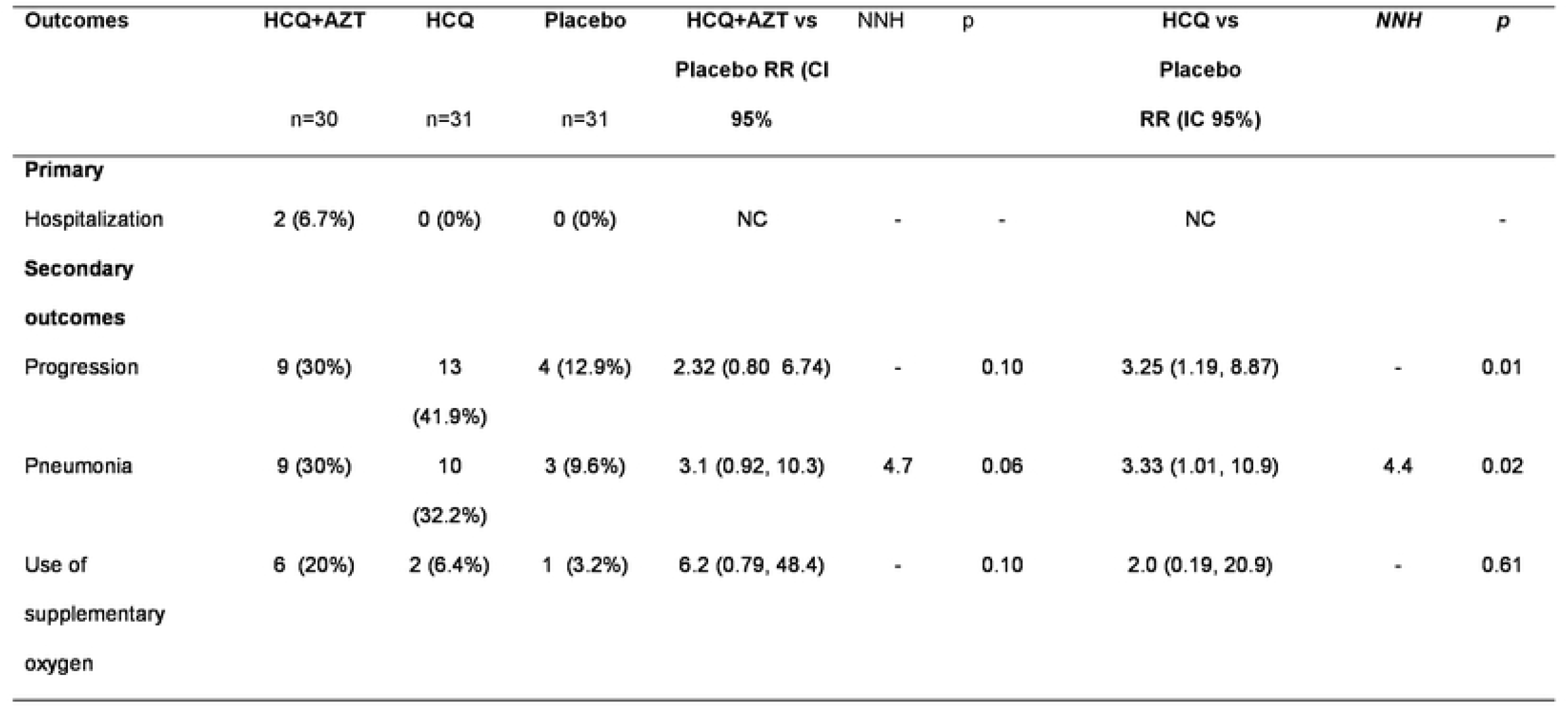
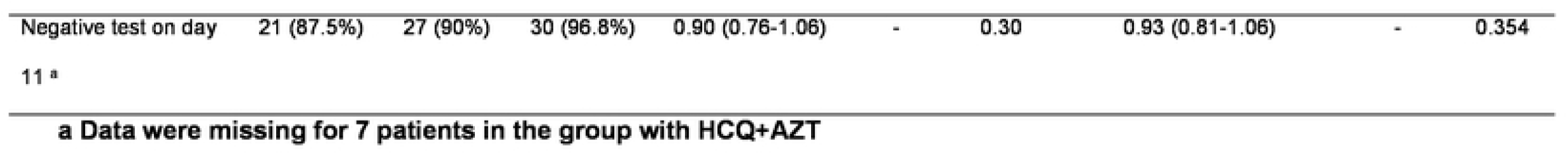
Efficacy of treatment for primary and secondary outcomes

### Secondary Outcomes

The incidence of disease progression was 30% (9/30) in the HCQ+AZT group, 41.9% (13/31) in the HCQ group, and 12.9% (4/31) in the placebo group. The RR for the HCQ+AZT group versus placebo was 2.32 (95% CI, 0.80-6.74; p=0.10) and the RR for the HCQ group versus placebo was 3.25 (95% CI, 1.19-8.87; p=0.01). Even though there was a statistically significant risk for developing pneumonia in the HCQ group compared to the placebo group [RR=3.33 (CI 95% 1.10, 10.9; p=0.02)], the pneumonia incidence was 30% (9/30) in the HCQ+AZT group, 32.2% (10/31) in the HCQ group and 9.6% (3/31) in the placebo group. The fixed combination of HCQ+AZT did not benefit compared to placebo. The frequency of patients who required supplemental oxygen was 20% (6/30) in the HCQ+ AZT group, 6.4% (2/31) in the HCQ group, and 3.2 % (1/31) in the placebo group. There were no significant differences between the groups. (Figure 1) There was no statistical difference between groups for negative test (PCR) on day 11.

Table 4 shows the efficacy of a placebo for secondary outcomes. For the group receiving placebo, we found disease progression in 12.9% (4/31) of participants compared to the group receiving active drugs (HCQ+AZT, HCQ) in 36.1% (22/61), [RR of 0.35 (0.13, 0.94; p=0.02)]. When analyzing pneumonia development, the results showed an incidence of 9.6% (3/31) compared to an incidence of 31% (19/61) in the group with HCQ+AZT, HCQ, [RR of 0.31 (0.09, 0.96, p=0.02)]. We could not observe differences in the use of supplemental oxygen.

**Table 4.** Efficacy of placebo vs active treatment for primary and secondary outcomes

| Outcomes | Placebo<br>n=31 | Otros tratamientos<br>n=61 | RR | p | NNT | RAR |
| --- | --- | --- | --- | --- | --- | --- |
| **Secondary** |  |  |  |  |  |  |
| Progression | 4 (12.9%) | 22 (36.1%) | 0.35 (0.13, 0.94) | 0.02 | 4.3 | 23.2 |
| Pneumonia | 3 (9.6%) | 19 (31%) | 0.31 (0.09, 0.96) | 0.02 | 4.6 | 21.4 |
| Use of supplementary<br>oxygen | 1 (3.2%) | 8 (13.1%) | 0.24 (0.03, 1.87) | 0.13 | - | - |

### Adverse Events

The most frequent adverse events were gastrointestinal symptoms with a rate of 16.6% (nausea, vomiting, heartburn, abdominal pain and distension), bradycardia with a rate of 6.4%, platelet elevation with a rate of 4.3%, and hypertriglyceridemia with a rate of 3.8%. Adverse events occurred with similar frequency between the different treatment groups. The analysis of whether there were differences in the QTc interval by the treatment group showed no differences in those who received HCQ+AZT or HCQ compared to the placebo group (Table 5).

**Table 5.** Adverse events according to treatment group

| Adverse event | Total | HCQ+AZT | HCQ | Placebo | *p* |
| --- | --- | --- | --- | --- | --- |
|  | 559 | 177 | 213 | 169 |  |
| Duration of the QTc segment (ms) at the end of the follow-up <sup>a</sup> | 416 (390- 434) | 413 (387-439) | 421 (396-430) | 413 (384-435) | 0.903 |
| Gastrointestinal, n (%) | 93 (16.6) | 33 (18.6) | 36 (16.9) | 24 (14.2) | 0.269 |
| Urticaria, n (%) | 5 (0.9) | 0 (0) | 4 (1.9) | 1 (0.6) | 0.541 |
| Dizziness, n (%) | 14 (2.5) | 3 (1.7) | 5 (2.3) | 6 (3.6) | 0.271 |
| Hypoglicemia, n (%) | 12 (2.1) | 3 (1.7) | 4 (1.9) | 5 (3.0) | 0.421 |
| Thrombocytosis, n (%) | 24 (4.3) | 6 (3.4) | 9 (4.2) | 9 (5.3) | 0.376 |
| Hypertriglyceridemia, n (%) | 21 (3.8) | 5 (2.8) | 8 (3.8) | 8 (4.7) | 0.508 |
| Bradycardia, n (%) | 36 (6.4) | 13 (7.3) | 11 (5.2) | 12 (7.1) | 0.916 |
| Others, n (%) | 349 (62.4) | 111 (62.4) | 134 (62.9) | 104 (61.5) | 0.573 |
| Median, interquartile range, Kruskal Wallis test |  |  |  |  |  |

## Discussion

At the beginning of the pandemic, the fixed combination of HCQ +AZT was proposed as a possible therapeutic option based on its antiviral and anti-inflammatory activity in vitro. However, clinical studies yielded inconsistent results. Therefore, this research project is conducted [18].

The study population consisted of participants with mild COVID-19, who had a low risk of complications. Of whom, only 2% required hospitalization. These data coincide with those reported in a population study from Alberta, Canada [19].

The results of this study suggest that the fixed combination of HCQ+AZT for the treatment of patients with mild COVID-19 does not reduce the risk of hospitalization compared to the use of HCQ alone or placebo. These results coincide with those reported in a Brazilian clinical trial on patients with mild and moderate COVID-19, which showed no significant differences for any of the treatment groups (HCQ+AZT, HCQ, or placebo) in terms of hospitalization or mechanical ventilation [20]. These results also coincide with those reported by other authors who evaluated the separate use of HCQ or AZT and found no benefit to reduce the risk of hospitalization, mechanical ventilation, or mortality [11,21–25].

The analysis of the secondary outcomes revealed that the administration of HCQ increased the risk of progression and pneumonia compared to placebo. However, for the combination of HCQ+AZT, the results showed only a trend in the risk for pneumonia, with no statistical significance, due to lack of statistical power. When we combined both active treatments against placebo for progression and pneumonia, we were able to show these differences.

The increase in the risk of damage after the use of the combination of HCQ+AZT, as in this study, was also reported by Kureder et al. who found a higher risk of mortality in patients who received this combination treatment, even in the multivariate model. However, these results were derived from an observational study where it was possible that the patients who received the use of HCQ+AZT had a more severe form of the disease at baseline, while in the present study, the participants included in the three treatment groups had mild symptoms [26].

Our results on the lack of benefit for the different outcomes of the study contrast with the results reported by Gautret et al., who highlighted the efficacy of the combination of HCQ+AZT for viral clearance on day six of treatment, in this study, no significant differences were found in the negativization of the PCR at day 11. It is possible to attribute the differences to the design of the study, non-randomization, the evaluation of an intermediate regulator as an outcome, and not evaluating other clinically relevant outcomes such as mechanical ventilation or mortality [27].

Although an antiviral effect against SARS-CoV-2 was found for HCQ and AZT in vitro, the use of HCQ in this study conditioned a higher risk of progression and pneumonia. This paradoxical response suggests that the drug could impair the immune response, conditioning a delay in the cellular and adaptive immune response, as shown by Roques et al. analyzing the effect of chloroquine on Chikungunya virus infection, who highlighted the contrast between the in vitro antiviral effect of chloroquine and the exacerbation of the disease in vivo and who reported that they are like the results reported by Maisonnasse, as well as our results [28,29].

Another concern about the use of HCQ+AZT for the treatment of COVID-19 infection is the risk of cardiovascular complications by prolonging the QT interval and causing polymorphic ventricular tachycardia in the form of Torsade de Pointes and death [30]. In this study, no differences in QT segment duration or fatal arrhythmias were documented in the three treatment groups, suggesting that significant cardiac involvement and arrhythmias in patients infected with COVID-19 have a multifactorial etiology and are more common in patients with severe forms of the disease, comorbidities, and advanced age [31, 32]. The absence of cardiovascular complications in our population can be explained by the inclusion of a young population, with a low prevalence of comorbidities and mild disease.

The most frequent adverse events were mild and gastrointestinal symptoms, characterized by nausea, vomiting, pain, and abdominal distension. The sample size calculation was 105 participants. However, the study was stopped after the inclusion of 92 participants due to a decrease in the number of patients in Mexico prior to the third pandemic wave. Participants who received ivermectin as concomitant treatment were included. However, the use or Ivermectin wasn’t associated with hospitalization, pneumonia and disease progression.

## Conclusion

The use of HCQ+AZT did not show efficacy in reducing the risk of hospitalization in patients with mild COVID-19 with controlled comorbidities. The use of HCQ and HCQ+AZT was associated with an increased risk of disease progression. The adverse events observed were mild and infrequent, predominantly consisting of gastrointestinal symptoms, without presenting cardiac alterations. Digital monitoring is a very valuable tool for the early detection of infection progression data. It is important to emphasize that empirical treatments were administered during the initial phases of the pandemic. It is important to continue generating evidence on this disease based on randomized control trials.

## Data Availability

All relevant data are within the manuscript and its Supporting Information files.

## Acknowledgments

This study is part of the doctoral studies at the Instituto Politécnico Nacional of the doctoral candidate Ivonne Anali Roy-Garcia.

We would like to thank all the patients who participate in this study, nevertheless all the requirements of the protocol. We also like to thank the directors of the Organ of Decentralized Administrative Operation Regional State of Mexico East and the directors of the Family Medicine Unit 52 Cuautitlán Izcalli and the Family Medicine Unit 28 for the facilities to carry out this research, Chief of Medical Benefit Services, Dr. María de los Ángeles Dichi Romero, Coordinator of Planning and Institutional Liaison, Dr. Olga Margarita Bertadillo Mendoza, Director of UMF 52, Dr. Irlanda León Bojorges and to the laboratory staff of the UMF 28 and the epidemiology, Dra. Brenda Jessica Espinosa Armenta.

